# How to improve adherence with quarantine: Rapid review of the evidence

**DOI:** 10.1101/2020.03.17.20037408

**Authors:** Rebecca K. Webster, Samantha K. Brooks, Louise E. Smith, Lisa Woodland, Simon Wessely, G. James Rubin

## Abstract

**Objectives:** The January 2020 outbreak of coronavirus has once again thrown the vexed issue of quarantine into the spotlight, with many countries asking their citizens to ‘self-isolate’ if they have potentially come into contact with the infection. However, adhering to quarantine is difficult. Decisions on how to apply quarantine should be based on the best available evidence to increase the likelihood of people adhering to protocols. We conducted a rapid review to identify factors associated with adherence to quarantine during infectious disease outbreaks.

**Study design:** Rapid evidence review.

**Methods:** We searched Medline, PsycINFO and Web of Science for published literature on the reasons for and factors associated with adherence to quarantine during an infectious disease outbreak.

**Results:** We found 3163 papers and included 14 in the review. Adherence to quarantine ranged from as little as 0 up to 92.8%. The main factors which influenced or were associated with adherence decisions were the knowledge people had about the disease and quarantine procedure, social norms, perceived benefits of quarantine and perceived risk of the disease, as well as practical issues such as running out of supplies or the financial consequences of being out of work.

**Conclusions:** People vary in their adherence to quarantine during infectious disease outbreaks. To improve this, public health officials should provide a timely, clear rationale for quarantine and information about protocols; emphasise social norms to encourage this altruistic behaviour; increase the perceived benefit that engaging in quarantine will have on public health; and ensure that sufficient supplies of food, medication and other essentials are provided.

## Introduction

Quarantine is the separation and restriction of movement of people who have potentially been exposed to a contagious disease, in order to limit disease spread. ^1^ This differs from isolation, which applies to people who have been diagnosed with the disease,^2^ although the terms are sometimes used interchangeably. Particularly during the early stages of a novel infectious disease outbreak, quarantine can be applied to large numbers of people. For example, in Toronto during the 2003 Severe Acute Respiratory Syndrome (SARS) outbreak, 100 people were placed into quarantine for every case that was diagnosed. ^3^ The early stages of the 2019 coronavirus outbreak have already witnessed the quarantining of entire cities within China, ^4^ while thousands of foreign nationals leaving China are being asked to enter quarantine at home or in government facilities upon return to their home countries.

The efficacy of quarantine is uncertain and in previous incidents its overuse has been criticised as lacking in scientific basis. ^3 5 6^ Regardless of this debate, one thing is clear: quarantine does not work if people do not adhere to it. While officially-sanctioned enforcement of quarantine orders is possible, ^7^ this can lead to legal dispute, ^5^ chaotic scenes of confrontation, ^8^ and poor mental health (which can occur even under voluntary procedures). ^4 9^ Many nations are understandably nervous of these outcomes, especially given that confrontation can now result in harrowing mobile phone footage making its way to social and mainstream media. In many societies it might also be difficult to persuade the police or military to forcibly prevent healthy people who wish to leave quarantine from doing so. Seeking to avoid instances of public backlash, many countries rely instead on a combination of inducements and appeals to civic duty in order to encourage people to adhere.

We present a rapid evidence review ^10^ of factors that increase or decrease adherence with quarantine requests.

## Methods

We used a search strategy including terms relating to quarantine (e.g. quarantine, patient isolation) and adherence (e.g. adherence, compliance). For the full search strategy, see Appendix 1.

Studies were eligible for inclusion if they a) reported on primary research; b) were published in peer-reviewed journals; c) were written in English, Italian or French (which could be translated by a member of our team); d) included participants asked to enter quarantine outside of a hospital environment for at least 24 hours; and e) included outcomes relating to factors associated with, or self-reported reasons for, adherence or non-adherence.

Two authors ran the search strategy on MEDLINE® on 27^th^ January 2020, and two authors ran the search strategy on PsycINFO and Web of Science on 30^th^ January 2020. Citations were downloaded to EndNote© version X9 (Thomson Reuters, New York, USA). The same authors who ran the search evaluated titles and abstracts, excluding any which were obviously irrelevant. We obtained full texts of remaining citations, and two authors reviewed these, excluding any which did not meet inclusion criteria. Finally, reference lists of remaining papers were hand-searched for additional relevant studies. We then compared results from full text screening; there were only minor discrepancies, which were resolved through discussion with the whole team.

The following data were extracted from included studies: authors, publication year, country of study, infectious disease outbreak, design and method, participants (including sample size and demographic information), reason for quarantine, length of quarantine, and key results. Data extraction was carried out by one author.

Narrative synthesis was used to analyse the results of the included papers and group results into related themes.

## Results

The initial search yielded 3163 papers, of which 14 included relevant data and were included in the review. Details of the screening stages can be seen in Figure 1. Characteristics of included studies and key results are presented in Table I. Nine studies reported adherence rates of quarantined individuals, which ranged from 0%-92.8%. We identified nine factors associated with adherence which are discussed below.

**Figure 1.**
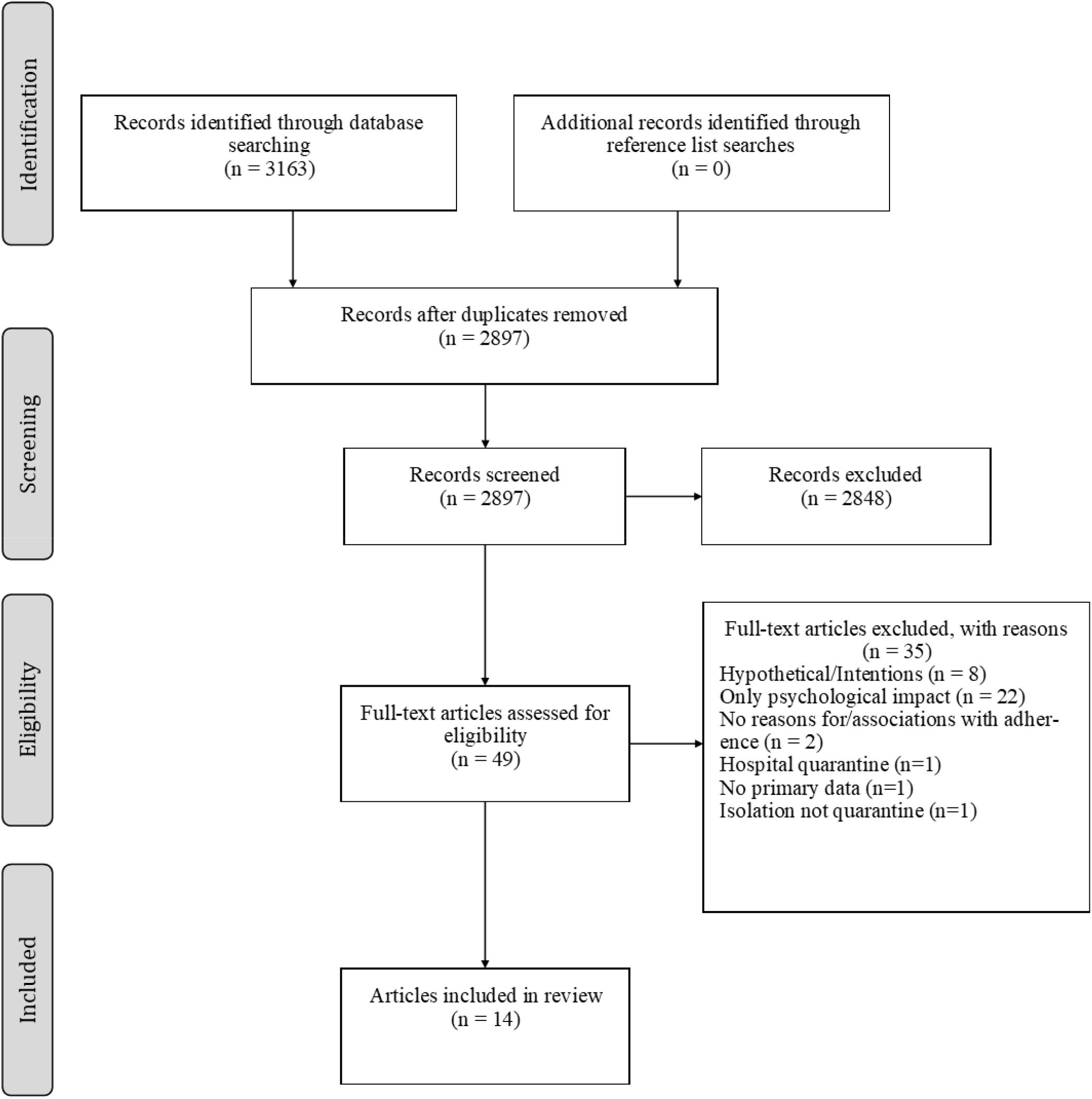
Flow diagram of screening results and reasons for exclusion.

**Table 1.** Characteristics of included studies and key results

| Study | Country | Disease | Design and method | Participants (N, Age, % male) | Quarantine protocol | Adherence rates | Factors associated with adherence |
| --- | --- | --- | --- | --- | --- | --- | --- |
| *Reasons given for adherence behaviour* |  |  |  |  |  |  |  |
| Braunack-mayer 2013 | Australia | Swine flu | Qualitative, interviews | School principals, staff, parents and students in five schools from an Australian city (56, -, -) | Home quarantine for seven days | - | Knowledge, Socio-cultural factors |
| Caleo 2008 | Sierra Leone | Ebola | Qualitative, semi structured face-to-face interviews | Households with and without Ebola cases and key community informants from a rural village (48, 18 (median), 47.3) | Restriction of movements during August 2014 | - | Knowledge, Socio-cultural factors, Perceived benefit of quarantine |
| Cava 2005 | Canada | SARS | Qualitative, semi structured face-to-face interviews | Individuals who had been quarantined (21, 18->65, 23.8) | Home quarantine for 10 days | “people adhered with differing levels of vigilance” | Socio-cultural factors, Perceived risk of disease, Practicalities |
| Desclaux 2017 | Senegal | Ebola | Qualitative, semi structured face-to-face interviews | Adult contact subjects and community volunteers (70, -, -) | Daily check-ups for physical symptoms with social distancing for 21 days | - | Knowledge, Socio-cultural factors, Perceived risk of disease, Trust in government |
| DiGiovanni 2004 | Canada | SARS | Qualitative, unstructured and structured face-to-face interviews, telephone polling, focus groups | Toronto residents affected by the SARS epidemic, affected by quarantine, and HCWs who had been quarantined (~1800, -, -) | Home quarantine for up to 10 days | - | Socio-cultural factors, Perceived benefit of quarantine, Practicalities, |
| Pellechia 2015 | Liberia | Ebola | Qualitative, focus groups and face-to-face semi-structured interviews | Individuals from 7 neighbourhoods and 5 villages (462, -, 60.6) | State enforced home and neighbourhood quarantine for 21 days | - | Practicalities |
| Teh 2012 | Australia | Swine flu | Quantitative, retrospective cohort study, telephone questionnaire | Participants tested for H1N1 and who were prescribed quarantine (538, -, -) | Home quarantine for seven days | 92.8% reported adherence to quarantine measures | Practicalities, Perceived risk of disease |

| *Factors tested for associations with adherence behaviour* |  |  |  |  |  |  |  |
| --- | --- | --- | --- | --- | --- | --- | --- |
| Hsu 2006 | Taiwan | SARS | Quantitative, cross-sectional paper questionnaire | HCWs in charge of SARS epidemic control at Health Centres in Taiwan (301, ≤30 - ≥50, -) | Home quarantine for 10-14 days | 0% - all nurses reported poor adherence from quarantined individuals | **Quarantine support characteristics, Knowledge, Perceived risk of disease** |
| Kavanagh 2011 | Australia | Swine flu | Quantitative, Cross-sectional online or telephone questionnaire | Parents from households with children who were placed in quarantine during the outbreak (297, -, 14.5) | Prescribed home quarantine for 1-14 days | 53% reported full adherence with quarantine within their household. | **Knowledge** |
| Kavanagh 2012 | Australia | Swine flu | Quantitative, Cross-sectional online or telephone questionnaire | Parents who were employed from households with children who were placed in quarantine during the outbreak (113, -, -) | Prescribed home quarantine for 1-14 days | Half of all households fully adhered with quarantine recommendations. | **Individual characteristics** |
| McVernon 2011 | Australia | Swine flu | Quantitative, Cross-sectional online or telephone questionnaire | Parents from households with children who were placed in quarantine during the outbreak (314, -, -) | Prescribed home quarantine for 1-14 days | 84.5% reported full adherence at household level | **Length of quarantine, Perceived risk of disease, Practicalities** |
| Porten 2006 | Germany | SARS | Quantitative, cross-sectional paper questionnaire | Respondents from local health departments (280, -, -) | Home quarantine for 10 days | - | **Individual characteristics** |
| Reynolds 2008 | Canada | SARS | Quantitative, cross-sectional paper questionnaire | Adults who were placed in quarantine (1057, 49.2, 37) | Prescribed home or work (for HCWs) quarantine for 2-10 days | 15.8% full adherence with all quarantine measures | **Individual characteristics, Perceived risk of disease** |
| Soud 2009 | United States | Mumps | Quantitative, cross-sectional telephone or face-to-face questionnaire | Students at a Kansas University with suspected mumps instructed to stay isolated (132, <20- ≥22, 37) | Prescribed home quarantine for 1-9 days | 75% stayed isolated for recommended number of days | **Length of quarantine, Perceived benefit of quarantine, Individual characteristics** |

Note: - , not reported, HCWs Health Care Workers
In many cases it was not clear how long participants were quarantine for. In these instances we have given the best estimates based on guidance by public health officials at that time

### Demographic and employment characteristics of those quarantined

There was mixed evidence as to whether demographic and employment characteristics of quarantined people affected adherence to quarantine protocol. Whether parents’ employers provided paid leave did not affect adherence to quarantine recommendations during the H1N1 outbreak among children who had been sent home from school. ^11^ However, where parents nonetheless took time off work to supervise their children, adherence to quarantine was higher, as the alternative might have involved others supervising children which would have broken quarantine protocol regarding social mixing. ^11^ Porten, et al. ^12^ found that during the SARS outbreak, unemployed or low-waged people were more likely to adhere to quarantine. For students, however, having an additional job alongside being a student did not appear to be a relevant factor. ^13^ Being a health care worker was associated with higher adherence to quarantine during the SARS outbreak in Canada. ^14^ Within student populations, no differences were found according to gender, age, full or part-time status, residing on or off campus, or quarantine location. ^13^

### Knowledge about the infectious disease outbreak and quarantine protocol

One of the major factors affecting adherence to quarantine is knowledge about the infection and the quarantine protocol. When five schools in an Australian city were closed during the H1N1 pandemic, a lack of clear quarantine instructions led some of those affected to invent their own quarantine rules, ^15^ seemingly based on what they thought constituted a visible symptom of the disease, the acceptable degree of contact with those infected and the risk of being affected or of infecting others. Parents in an Australian city who understood what they were meant to do during the quarantine period for H1N1 had significantly higher adherence to quarantine. ^16^ Caleo, et al. ^17^ found that people in Sierra Leone who were put under quarantine due to Ebola also had problems adhering to protocols because they did not understand what ‘isolation’ meant. Adherence to quarantine in Taiwan during the SARS outbreak was significantly associated with higher awareness of the pandemic. ^18^

However, in some cases, too much perceived knowledge might be a hindrance. Residents of villages that were quarantined during the Ebola epidemic who were health professionals often had more knowledge about Ebola than the volunteers sent in to support the village. They believed they knew more about the risk of infection than volunteers, but unlike the latter did not always adhere to the quarantine measures as they thought the restrictions were too over-precautionary. ^19^

One study looked at the effect of where people got their knowledge of quarantine protocols from, finding no difference in adherence rates between those that sourced information from official vs unofficial sources. ^16^

### Socio-cultural factors: social norms, cultural values and the law

Social norms play an important part in adherence to quarantine protocols. Many individuals quarantined during the SARS outbreak in Canada reported social pressure from others to adhere to quarantine. ^20^ Desclaux, et al. ^19^ noted that residents from villages in Senegal which quarantined during Ebola said that if there was favourable opinion for engaging in quarantine from the head of household, it was expected the rest of the household would follow suit and adhere. Residents also acknowledged a respect for the collective commitment to protect the community against Ebola which they did not want to be seen to be disrespecting.

However, social norms can also reduce adherence to quarantine. As rumours that others were breaking quarantine began to surface among Australian school communities quarantined during the H1N1 outbreak in Australia, those affected explained they were more likely to break quarantine protocols themselves. ^15^ Volunteers who were supporting villages in Senegal during quarantine for Ebola also mentioned ‘relaxing their principles’ and allowing non-adherence to quarantine at certain times in order to avoid direct challenges to containment which would then be seen by the rest of the village. ^19^

Cultural values also play an important part in decisions to adhere to quarantine. Residents of villages in West Africa quarantined during an Ebola outbreak often did not adhere to quarantine as it was inherent in their culture to care for people when they are sick, rather than ‘abandon’ them. ^17^ Conversely, two studies noted that participants quarantined during SARS explained that they adhered to quarantine as it was their ‘civic duty’ and they wanted to be a good citizen. ^20 21^

Two studies noted that ‘following the law’ was a reason for adhering to quarantine during the Ebola outbreak in Sierra Leone, ^17^ and the SARS outbreak in Canada. ^20^ In these circumstances, if individuals were found breaking quarantine rules, they faced paying fines. Relatedly, where the term ‘voluntary’ was used to describe quarantine in Canada during the SARS outbreak, residents correctly understood this meant that adherence was at their discretion, rather than enforced by the government, something which then reduced adherence. ^21^

### Perceived benefit of quarantine

People who perceive a benefit of quarantine are more likely to adhere to it. For example, as village residents began to notice a slowing in the spread of Ebola, their attitudes changed and adherence to quarantine protocols increased. ^17^ Toronto residents affected by quarantine for SARS explained they adhered to protocols because they believed this would reduce the risk of transmission to others. ^21^ Similarly, Soud, et al. ^13^ found that perceived higher importance of avoiding others during isolation was associated with adherence to quarantine during a mumps outbreak at a university in the United States.

### Perceived risk of the disease outbreak

People who perceive a disease outbreak to be riskier (in terms of disease transmission and severity of disease outcomes) are more likely to adhere to quarantine. Cava, et al. ^20^ found that those who adhered to quarantine for SARS had higher perceptions of risk for the disease. Residents in Senegalese villages quarantined due to Ebola adhered because they thought transmission could happen even when asymptomatic. ^19^ Higher perceived fear of SARS was associated with adherence to quarantine measures in Taiwan. ^18^ Conversely, reasons for non-adherence to quarantine in Australia during the H1N1 pandemic included belief that the disease was not serious. ^22^ When comparing quarantine adherence during two separate outbreaks of SARS in Canada, adherence was higher during the second outbreak. ^14^ Indeed this may be due to the second outbreak increasing the perceived severity of the outbreak as it had not receded, or it could be due to people being more knowledgeable about the disease and quarantine protocol the second time around. Relatedly, increased adherence to quarantine in Australia during the H1N1 pandemic occurred when there was an influenza case in the household, which again may be associated with increased perceived risk of disease transmission now that the disease is amongst family members, or an increase in knowledge of the disease and quarantine protocol. ^23^

One study looked at the effect of the objective severity of disease on adherence to quarantine, finding no effect of the total probable cases of SARS or number of quarantined people on likelihood of adherence. ^18^

### Practicalities of quarantine

Two studies reported the need to work and fear of loss of income as reasons for not adhering to quarantine protocols. ^21 22^ In Teh, et al. ^22^, participants also mentioned factors relating to ‘life carrying on’ outside of quarantine as reasons for not adhering. Examples included needing to attend an important event or visiting family and friends.

Three studies reported that participants needed to break quarantine protocol in order to get supplies ^21 22 24^ or to seek medical attention. ^22^

Sometimes factors relating to the household situation during quarantine influenced adherence. This could be due to people being preoccupied with the ill health of a loved one, such that they did not adhere to quarantine protocols themselves. ^20^ Similarly, if quarantined children were able to be cared for by adults within the household rather than by outside family, friends or hired help coming to the house, this made it easier for families to adhere to quarantine protocol.^23^

### Experience and belief of healthcare workers and functioning of health centres

There was no evidence of healthcare workers’ experience or beliefs surrounding the outbreak affecting adherence to quarantine protocol. Hsu, et al. ^18^ found no effect of healthcare workers years of experience or perceived severity of the epidemic on individuals’ adherence to quarantine protocol during the SARS epidemic in Taiwan. However, there was some evidence that the good functioning of health centres in Taiwan that were helping to control the SARS outbreak were associated with increased adherence. Hsu, et al. ^18^ found that if health centres were functioning well and received adequate resourcing, this was associated with increased adherence by people in quarantine. What did not seem to influence adherence was whether the help came from volunteers or trained staff.

### Length of quarantine

There was mixed evidence for whether the length of prescribed quarantine affected adherence to quarantine protocol. There was no effect of the length of prescribed quarantine for households during the H1N1 pandemic in Australia ^23^. Conversely, a quarantine duration of one to four days was associated with higher adherence than a duration of five to nine days during a mumps outbreak at an American University. ^13^

### Trust in government

People in Senegal who had a pre-existing positive appraisal of the health care system and had trust in the national response to Ebola were more likely to adhere to quarantine. ^19^

## Discussion

Although the effectiveness of quarantine is not always clear-cut ^3 5 6^, if public health officials deem it is necessary then it is important to understand how to encourage people to adhere to quarantine protocols. Our review found that adherence to quarantine during infectious disease outbreaks can be variable. In the studies we reviewed, adherence ranged from 0 to 93%. The most common factors affecting people’s adherence to quarantine were their knowledge about the infectious disease outbreak and quarantine protocol, social norms, perceived benefits of quarantine, perceived risk of disease and practicalities of being in quarantine. These factors have also been found to influence adherence to other protective health behaviours with regards to infectious diseases such as handwashing, wearing face masks, avoiding crowds and vaccination. ^25 26^ The recommended actions for increasing adherence to voluntary quarantine are discussed below, and a summary of key points is shown in Figure 2

**Figure 2.**
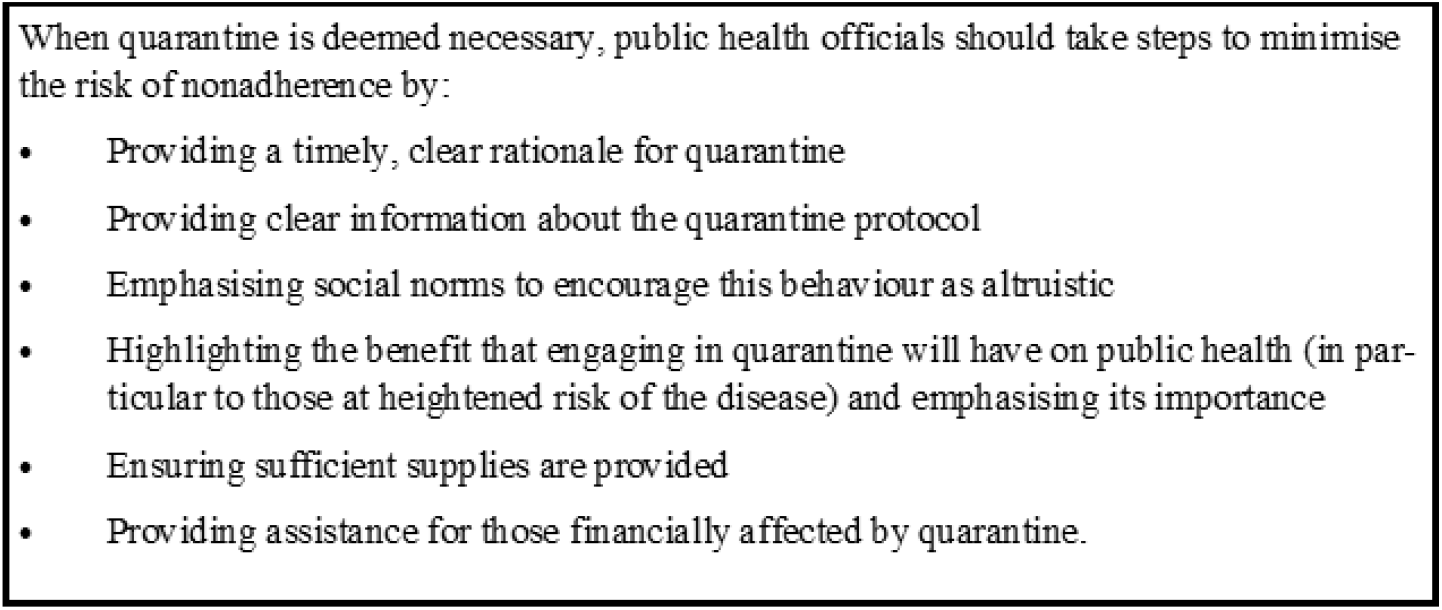
Summary of key recommendations.

As compulsory quarantine on any large scale is almost certainly not practicable in a democratic society, public health officials must do everything they can to encourage voluntary adherence to quarantine protocols. Key to this is making sure that information about the infectious disease outbreak and quarantine protocol is clear and consistent. Where information is unclear and open to interpretation, this can lead to people creating their own, possibly ineffective, rules. ^15^ In the era of ‘fake news’ and rumour we appreciate consistent messaging is difficult, but it remains the case that leaving the information needs of the public unmet can be dangerous. Public health teams should regularly check with those under quarantine what they understand or are unclear on, and provide clear, authoritative information where needed.

It is also important to reinforce social norms and moral values around quarantine. These are recognised determinants of behaviour. ^27^ Many participants included in our reviewed studies reported social pressure from others to comply, ^20^ not wanting to be seen going against the collective commitment to protect against the outbreak, ^19^ and feeling quarantine was their ‘civic duty’.^20 21^ Emphasising the altruistic nature of engaging in quarantine may help promote these beliefs.

It is likely, however, that appeals to altruism would be quickly undermined if practical or logistical problems began to appear. Fear of losing income, running out of supplies, lack of staff and related issues must be anticipated and prevented. ^18 21 22 24^ Care must also be taken to monitor, and intervene in, emerging social norms that may not support quarantine, for example rumours of others breaking quarantine without apparent detrimental effect. ^15 28^ At the same time the public need to be assured why quarantine is necessary (focussing on the perceived risks of the disease ^13 14 18^-^20 23 29^) and that it is important for everyone affected to engage with it. As with other health behaviours, ^30 31^ as perceptions of the benefit of quarantine increase, so too should adherence. ^13 17 21^

### Strengths and limitations

Given the rapid and evolving nature of the coronavirus outbreak and the need for guidance to support quarantine efforts, this rapid review was limited to peer-reviewed publications of primary data without searching grey literature and did not include a formal quality assessment of included studies. As such it important to note the review is not exhaustive and may have missed key articles in the search results and relevant articles may have been excluded as they were published in languages other than English, Italian and French. In addition, readers should read our interpretations of the evidence with caution as the quality of the studies is not known. We did, however, search reference lists to identify papers that may not have been found in the initial search and engaged multiple members of the team in the screening process to improve methodological rigour.

Our recommendations are primarily based on results from studies of small groups of people in home quarantine due to a small selection of infectious disease outbreaks in a limited number of countries. Whilst we anticipate that many of the risk factors for adherence would likely be similar for larger quarantine approaches, such as for whole towns or cities, and for other types of infectious disease outbreaks, there are also likely to be differences in such situations that mean the recommendations presented in this paper should only be applied to such situations cautiously. However, while this review cannot provide recommendations that will encourage adherence in every future quarantined population, the lessons from our review may be a good starting point for those considering these situations.

### Conclusion

People vary in their adherence to quarantine during infectious disease outbreaks. Adherence depends on the psychological and practical factors associated with infectious disease outbreaks and quarantine. When quarantine is deemed necessary, public health officials should take should steps to minimise the risk of non-adherence by providing a timely, clear rationale for quarantine and information about protocols; emphasising social norms to encourage this altruistic behaviour; increasing the perceived benefit that engaging in quarantine will have on public health (in particular to those at heightened risk of the disease); and ensuring sufficient supplies are provided.

## Data Availability

None available. This is a rapid review and not based on original data

## Appendix I

### Search strategy

Medline:

1. quarantine.mp. or exp Quarantine/
2. exp Patient Isolation/
3. psych*.mp.
4. exp Social Stigma/ or stigma.mp.
5. adheren*.mp.
6. complian*.mp.
7. 1 or 2
8. 3 or 4 or 5 or 6
9. 7 and 8

PsycInfo/Web of Science:

1. exp Social Isolation/ or quarantine.mp.
2. (isolation and (infect* or SARS or influenza or flu or MERS or ebola)).mp.
3. psych*.mp.
4. stigma.mp. or exp Stigma/
5. adheren*.mp.
6. complian*.mp.
7. 1 or 2
8. 3 or 4 or 5 or 6
9. 7 and 8

